# Development and validation of a clinical scoring system to predict left ventricular systolic dysfunction in patients with left bundle branch block

**DOI:** 10.1101/2025.05.08.25327272

**Authors:** Chun Kai Chen, Yen Bin Liu, Chu Chih Chen, Kuo Liong Chien, Hui Chun Huang

**Affiliations:** Department of Internal Medicine, Division of Cardiology, National Taiwan University Hospital and National Taiwan University College of Medicine, Taipei, Taiwan; Institute of Epidemiology and Preventive Medicine, College of Public Health, National Taiwan University, Taipei, Taiwan; Division of Biostatistics and Bioinformatics, Institute of Population Health Sciences, National Health Research Institutes, Taiwan; Research Center for Environmental Medicine, Kaohsiung Medical University, Taiwan

**Keywords:** left bundle branch block, reduced systolic function, heart failure, prediction

## Abstract

**Background:** Left bundle branch block (LBBB) adversely affects left ventricular (LV) synchrony and function, resulting in poorer outcomes for patients with systolic heart failure or coronary artery disease (CAD). Recently, LBBB-induced cardiomyopathy was recognized as a potentially reversible condition. Nevertheless, there is a significant knowledge gap regarding how to identify patients with LBBB at risk of reduced LV systolic function, leading to uncertainties in screening strategies. This study aimed to develop and validate a prediction model for detecting reduced LV systolic function among patients with LBBB morphology on electrocardiogram.

**Methods:** This study adhered to the Transparent Reporting of a Multivariable Prediction Model for Individual Prognosis or Diagnosis reporting guidelines. We retrospectively enrolled patients with LBBB morphology from the National Taiwan University Hospital between 2010 and 2018. Patients were randomly divided into development and validation sets. Predictors of reduced LV systolic function were identified from medical histories and electrocardiogram features, and a multivariable logistic regression model was developed.

**Results:** We enrolled 821 patients with LBBB morphology, divided into development (411 patients) and validation cohorts (410 patients). The final predictive model incorporated several variables, including younger age, male sex, hypertension, CAD, history of congestive heart failure, and discordant LBBB. The model showed good discriminative power with a C-statistic of 0.816 and a bias-corrected C-statistic of 0.807, indicating adequate calibration. Additionally, a clinical scoring system named CCDS65 was developed to categorize patients into groups based on their predicted risk of reduced left ventricular systolic function.

**Conclusions:** We developed and internally validated a straightforward predictive model to identify reduced LV systolic function in patients with LBBB morphology simply through history taking and ECG morphology. The CCDS65 scoring system provides a practical tool for quickly detecting reduced LV systolic function in clinical practice, enabling timely management through guideline-directed medical therapy and pacing interventions.

**Clinical Perspective:**

- **What is new?**

We successfully developed the CCDS65 scoring system, which effectively predicts echocardiographic reduced left ventricular systolic function in patients with complete LBBB. The predictive model incorporated several variables, including younger age, male sex, hypertension, history of coronary artery disease (CAD), history of congestive heart failure (HF), and discordant LBBB.

- **What are the clinical implications**?

Given the advancements in guideline-directed medical therapies and left bundle branch area pacing, early detection of reduced LV systolic function in patients with complete LBBB is increasingly important. The CCDS65 scoring system provides a practical tool for quickly detecting reduced LV systolic function in clinical practice, enabling timely management through guideline-directed medical therapy and pacing interventions.

## Introduction

The occurrence of left bundle branch block (LBBB) adversely affects left ventricular (LV) mechanical synchrony and can negatively impact LV function. LBBB is linked with poorer outcomes in patients with systolic heart failure (HF) or coronary artery disease (CAD) (1).Recently, LBBB-induced cardiomyopathy has been acknowledged as a reversible form of cardiomyopathy (2,3). Effective detection and management of this condition can lead to the restoration of LV ejection fraction (LVEF) and functional class and potentially reduce morbidity and mortality (3). Despite these advancements, a significant knowledge gap remains in predicting patients with LBBB with a risk of reduced LV systolic function, leading to an unclear screening strategy for patients with this condition.

This study aimed to develop and validate a predictive model designed to detect reduced LV systolic function within 6 months before or after the index electrocardiogram (ECG) in patients with complete LBBB morphology.

## Methods

The methodology of this study, aimed at generating a predictive model, adheres to the Transparent Reporting of a Multivariable Prediction Model for Individual Prognosis or Diagnosis reporting guidelines (4). The study protocol was approved by the National Taiwan University Hospital (NTUH) Research Ethics Committee (Study No. 202501026MIND), which also waived the requirement for informed consent. This study complies with the principles of the Declaration of Helsinki.

### Patient selection

We retrospectively enrolled all patients with LBBB morphology on ECG at NTUH and its branches from January 2010 to December 2018. The definition of LBBB adhered to the criteria set by the American Heart Association, the American College of Cardiology Foundation, and the Heart Rhythm Society for diagnosing cardiac conduction disturbances (5). A total of 1,835 adult patients with LBBB morphology on ECG were initially enrolled. Baseline characteristics were obtained from the medical records, and serial follow-up echocardiograms were tracked and documented. Patients with pacing rhythm, transient LBBB, acute events such as acute coronary syndrome within 6 months, and those without baseline transthoracic echocardiography results within 6 months of ECG were also excluded., leaving 821 patients in the final analysis (Figure 1).

**Figure 1:**
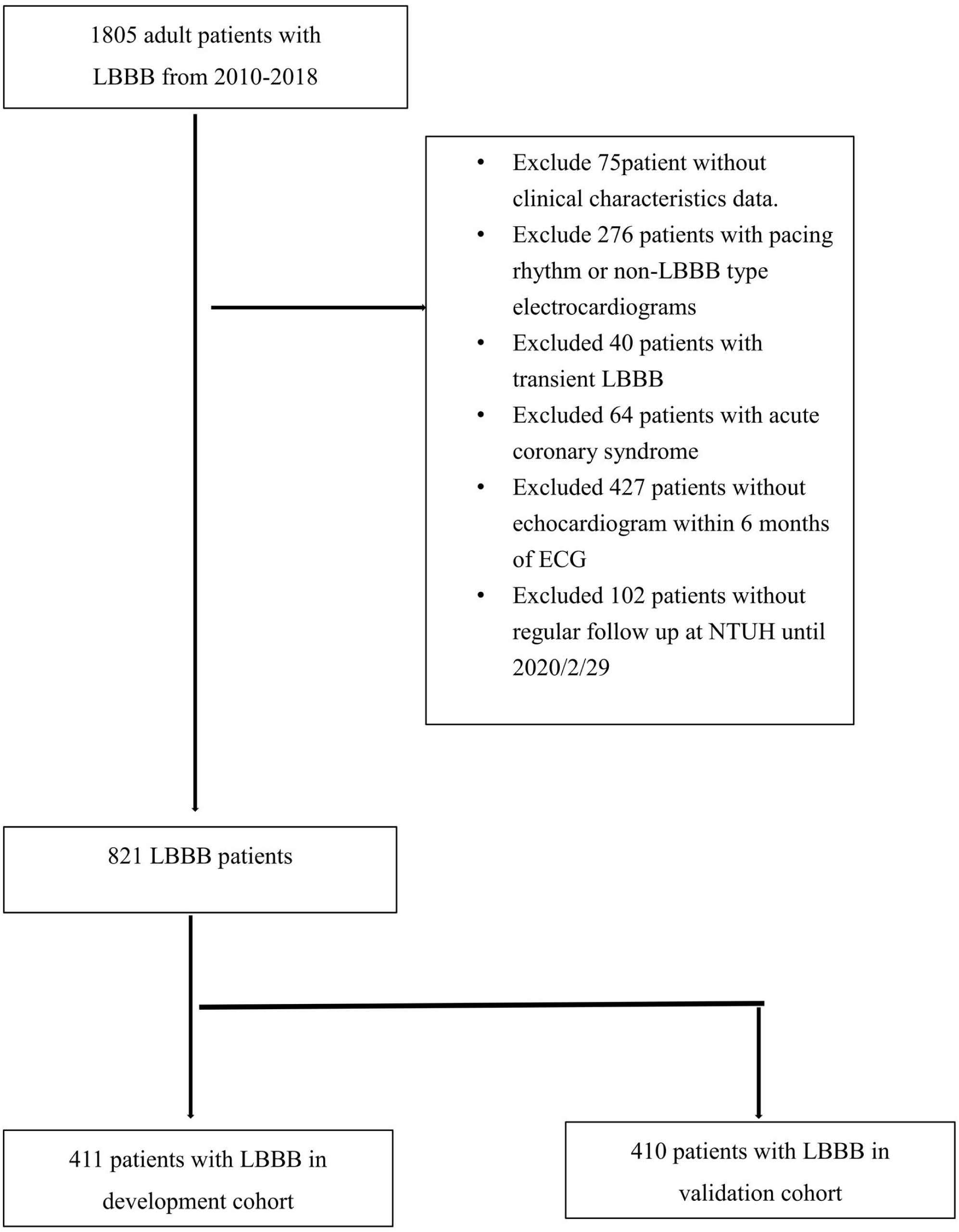
Patient selection flow chart. EF, ejection fraction; LBBB, left bundle branch block; NTUH, National Taiwan University Hospital.

### Development and validation cohorts

The included population was randomly divided into two cohorts based on the LVEF assessed by echocardiogram. The first cohort served as the basis for developing the prediction model (the development cohort), while the second was used for model validation (the validation cohort) (Figure 1).

### LVEF measurement

We tracked and obtained echocardiograms from patients within 6 months before or after the date the index ECG. These echocardiogram images were acquired following the guidelines recommended by the American Society of Echocardiography (6). We used the Teichholz method to measure various cardiac dimensions, including the interventricular septal diameter, left ventricular end-diastolic diameter, left ventricular posterior wall diameter, left ventricular end-systolic diameter, and left ventricular ejection fraction. For patients with LBBB, LVEF was measured using apical four-chamber and two-chamber views by the Simpson’s biplane method.

### Primary outcome

The primary outcome was reduced left ventricular systolic function, defined as LVEF of <40%. Patients in each cohort were randomly divided into groups with normal LVEF and reduced left ventricular systolic function at a ratio of 2 to 1 (Figure 1).

### Predictors

We conducted within-group comparisons to identify statistically significant characteristics among the patients. These characteristics were then used as predictor variables for reduced left ventricular systolic function.

### Data collection and handling missing data

We accessed the medical record database at NTUH to gather comprehensive data. Collected parameters included age, sex, body mass index (BMI), CAD risk factors, chronic obstructive pulmonary disease (COPD), cerebral vascular events, histories of heart failure, renal insufficiency, and atrial fibrillation. Each patient had an index ECG from which we recorded heart rate, QRS duration, QTc interval, left axis deviation, left atrial enlargement, left ventricular hypertrophy, and T wave concordance/discordance at leads I, V5, and V6.

## Statistical analysis

### Patients’ baseline characteristics and electrocardiogram and echocardiogram data

Categorical variables are reported as numbers (percentages), while continuous variables are presented as either means ± standard deviation (SD) for those with a normal distribution or medians (interquartile ranges) for non-normal distributions. To facilitate the statistical analysis, logarithmic transformation was applied to non-normally distributed variables to approximate normal distribution. Statistical significance across different groups was assessed using the appropriate tests: Student’s t-test for normally distributed continuous variables and nonparametric tests—either the Wilcoxon rank-sum test or the Kruskal−Wallis test—for those without normal distribution. Categorical variables were analyzed using Fisher’s exact test or the chi-squared test, depending on suitability. All statistical analyses were performed using SAS/STAT version 15.2 for Windows and R version 4.4.3, employing the rms package for enhanced methodology.

### Development of the prediction model

In the development cohort, we identified parameters that showed statistically significant differences between groups and considered them as potential predictors. We initially calculated crude odds ratios with 95% confidence intervals for these predictors using univariate logistic regression. Parameters with a significance level of p < 0.01 in the univariate analysis were selected for model creation. Subsequently, we employed a multivariable logistic regression model to determine the β-coefficients with standard errors for each selected variable. Model performance and internal validation were evaluated using several metrics: Somers’ D correlation, C-statistics, Nagelkerke R^2^ value, calibration intercept and slope, and Brier score (Supplementary Table 2). Additionally, we conducted a Hosmer−Lemeshow goodness-of-fit test to examine calibration within the development cohort. Calibration plots, using locally weighted scatterplot smoothing, were created to visually represent the relationship between predicted and observed outcomes (Figure 2). Internal validation of the model was further supported by a bootstrapping procedure involving 200 samples drawn with replacement from the original dataset.

**Figure 2:**
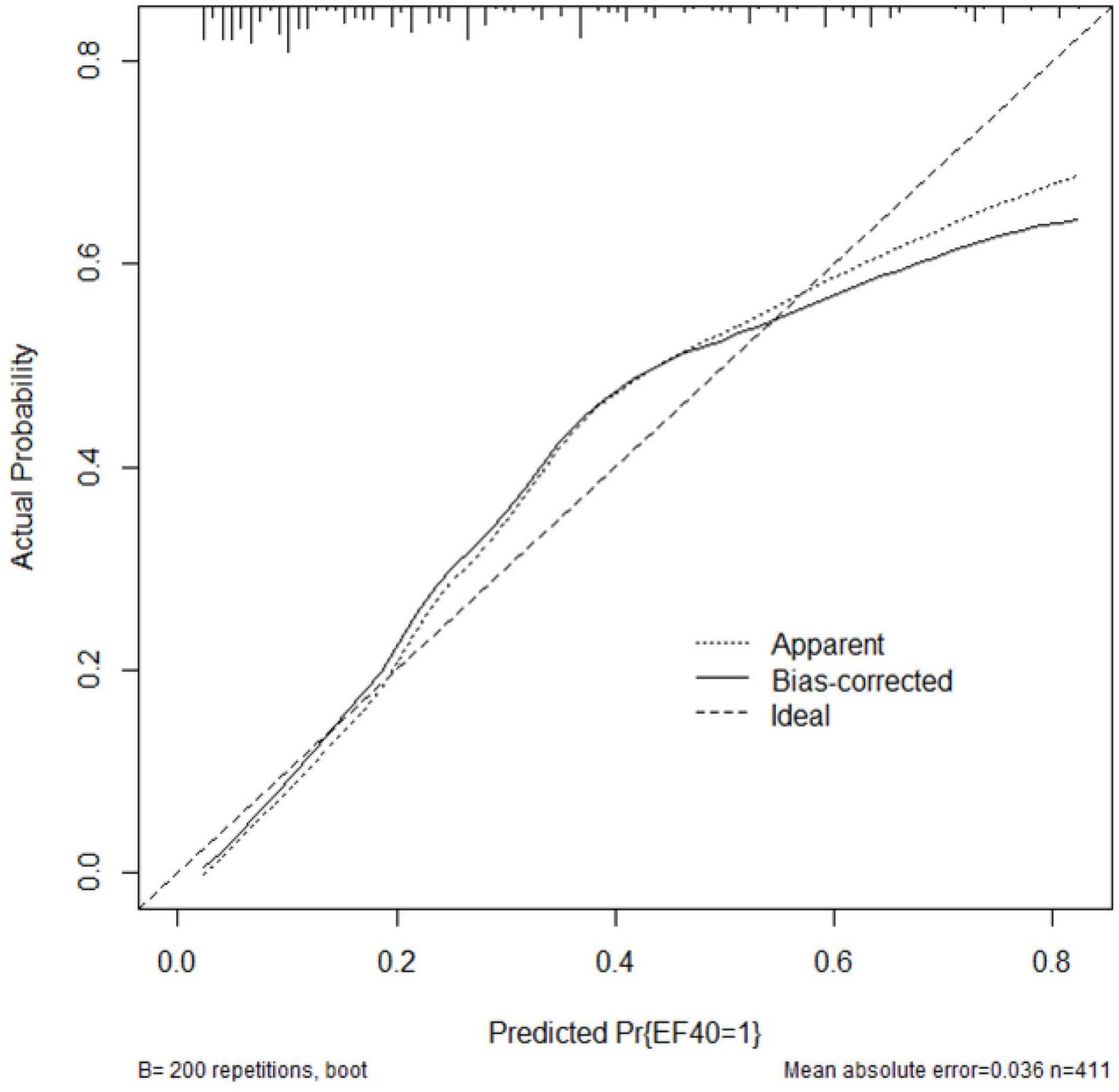
Calibration plot of the model in the development cohort. X-axis: Predicted probability, Y-axis: Observed, Apparent: original model, Bias-corrected: bias corrected by bootstrapping. Original prediction model is evaluated by bootstrapping (n=200) in development cohort. This model is described in Table 2 of the main text

**Table 1.**
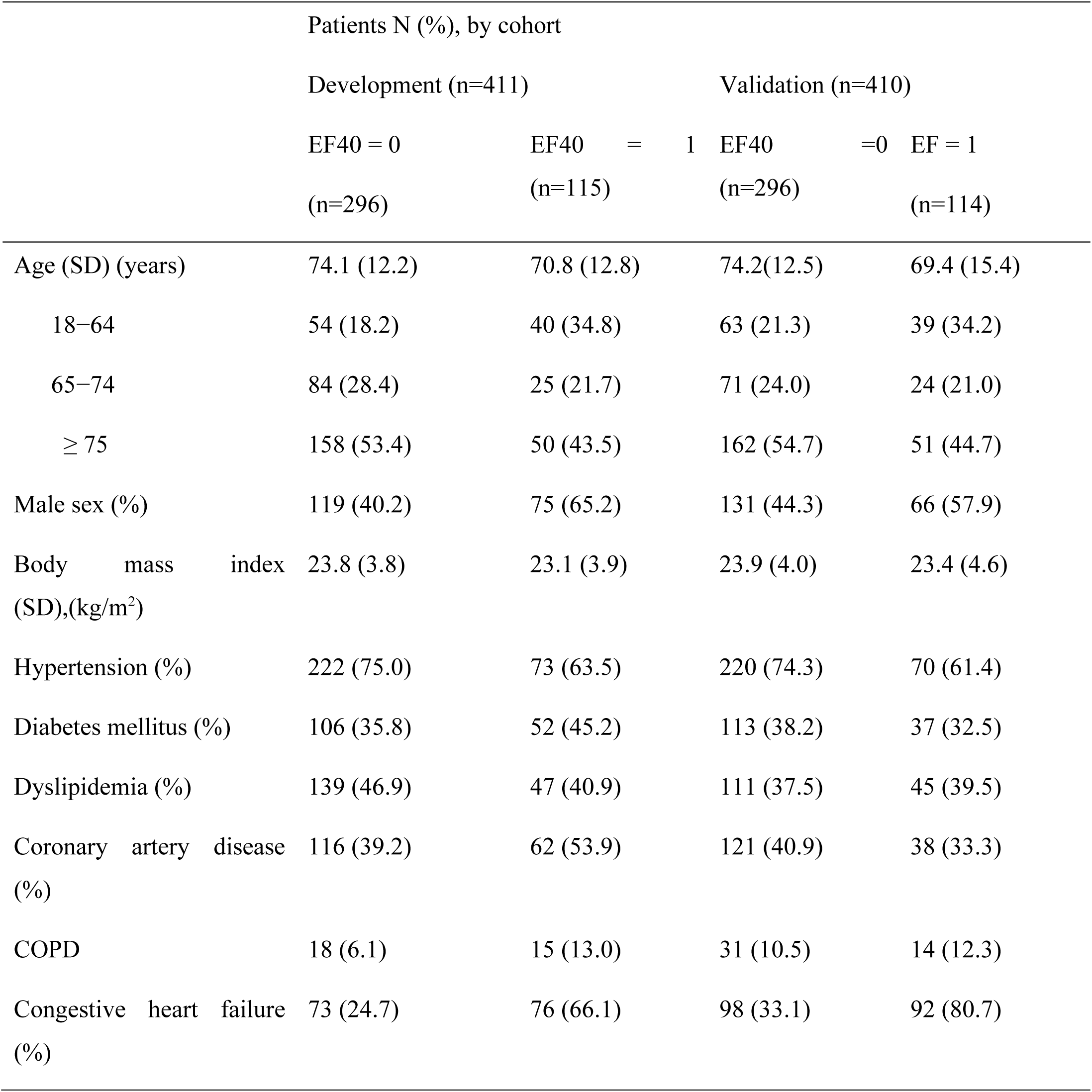
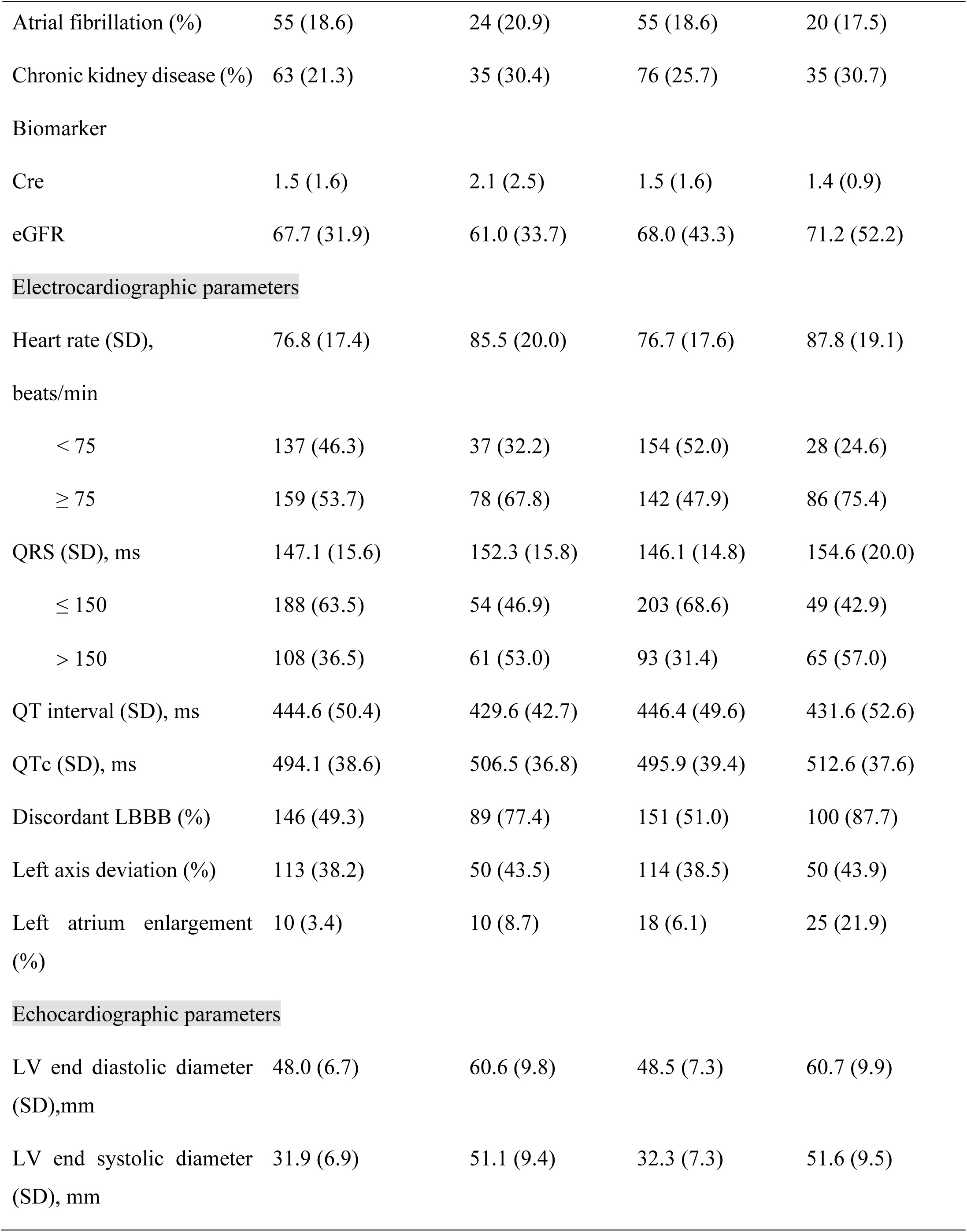
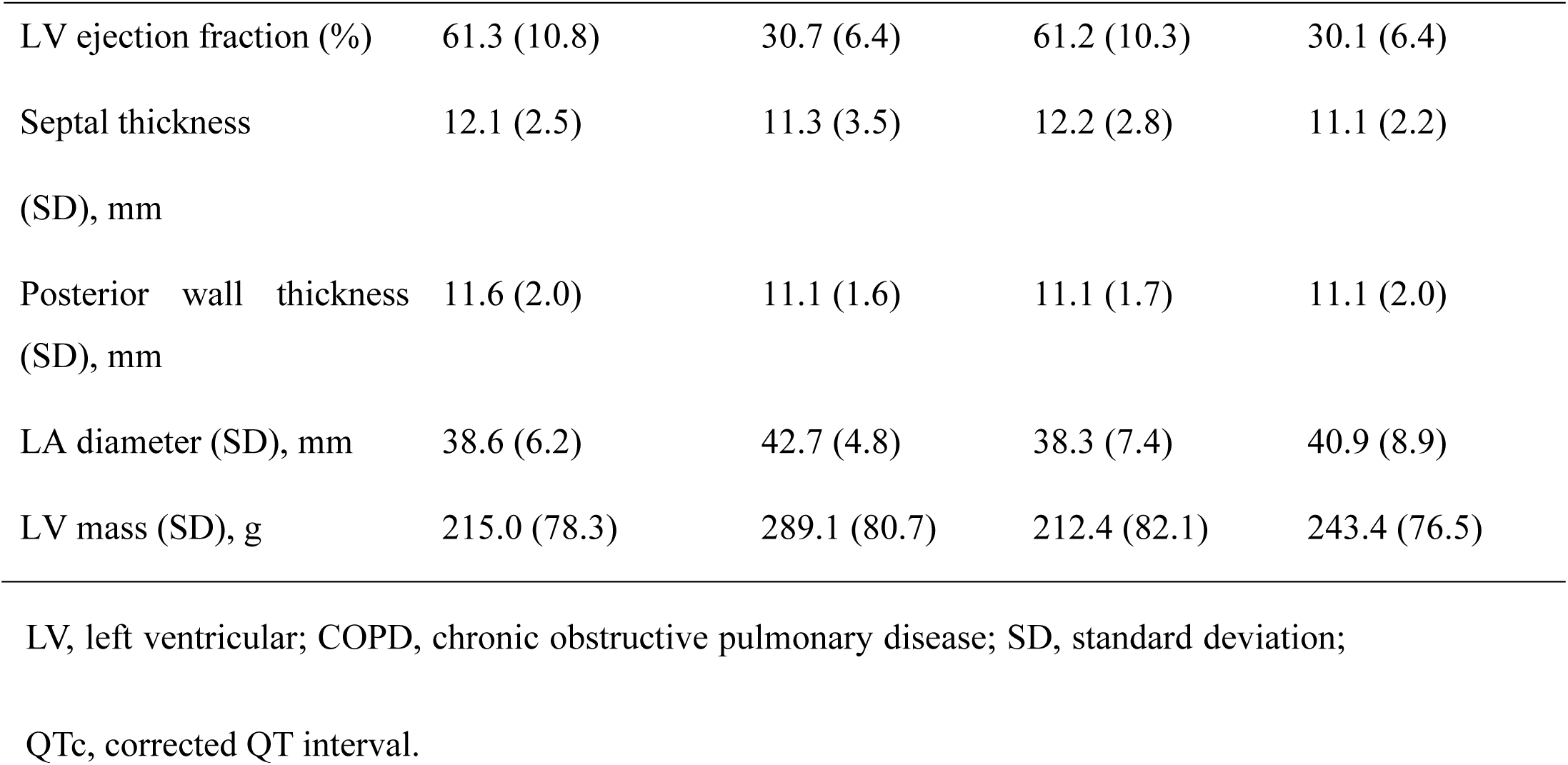
Baseline demographic and clinical characteristics and electrocardiographic parameters of patients with LBBB.

**Table 2.** Multivariate logistic model of the primary endpoint (EF<40%) after adjusting for clinical characteristics and electrocardiography and echocardiography parameters.

|  | Odds |  |  |  |  |
| --- | --- | --- | --- | --- | --- |
|  | coef | (SE) | ratio | 95% CI | p-value |
| Intercept | -4.43 | (0.56) |  |  | **<.0001** |
| Age, 18–64 years | 0.95 | (0.29) | 2.58 | 1.46–4.54 | **0.001** |
| Sex, males | 0.89 | (0.26) | 2.44 | 1.46–4.07 | **0.0006** |
| History of CAD | 0.56 | (0.26) | 1.75 | 1.06–2.89 | **0.03** |
| History of CHF | 1.73 | (0.26) | 5.64 | 3.39–9.39 | **<.0001** |
| Discordant LBBB | 1.05 | (0.28) | 2.84 | 1.64–4.92 | **0.0002** |
| Heart Rate, $\geq 75$ | 1.65 | (0.27) | 1.65 | 0.97-2.80 | 0.06 |
| QRS duration | 1.18 | (0.27) | 1.18 | 0.70-1.99 | 0.52 |

CHF, congestive heart failure; LBBB, left bundle branch block; CAD, coronary artery disease; CI, confidence interval

### Scoring and grouping

A scoring system was established based on the β-coefficients of each variable from the development cohort, assigning a simple integer score to predict the outcome. The diagnostic capabilities of each score were assessed, including the sensitivity, specificity, positive likelihood ratio, negative likelihood ratio, as well as rates of true positives, true negatives, false positives, and false negatives. Subsequently, patients were stratified into four distinct groups according to their scores.

### Validation of the prediction model, scoring, and grouping

The model developed in the initial cohort was subsequently applied to the validation cohort to test its efficacy. The performance of the prediction model was rigorously evaluated to determine its accuracy in predicting outcomes. Calibration was performed to assess the alignment between the predicted outcomes and the actual observed outcomes across each of the defined groups. This step ensured the model’s reliability and applicability in a clinical setting.

## Results

### Study population

Utilizing data from the NTUH database, we enrolled 821 patients who met the study’s eligibility criteria. The participants were allocated into two cohorts: 411 in the development cohort and 410 in the validation cohort. In these groups, 115 and 114 patients, respectively, developed reduced LV systolic function (Figure 1). In the development cohort, statistically significant factors associated with reduced LV systolic function included younger age, male sex, lower incidence of hypertension, history of CAD, COPD/asthma, history of CHF, higher heart rate, prolonged QRS duration, extended QTc interval, elevated creatinine levels, and the presence of discordant LBBB (Table 1).

### Model development

We selected factors associated with reduced LV systolic function as predictor candidates, including diabetes mellitus, dyslipidemia, cerebral vascular events, atrial fibrillation, and renal insufficiency, based on established links with heart failure. Crude odds ratios (OR) with 95% confidence intervals (CI) for these candidates were calculated using univariate logistic regression (Supplementary Table 1). Predictors with p<0.01 were chosen for model development. These included age between 18 to 64 years, male sex, hypertension, history of CAD, history of CHF, heart rate ≥ 75 bpm, QRS duration > 150 ms, and the presence of discordant LBBB. These variables were included in a multivariate logistic regression model, where the β-coefficient and standard error (SE) were calculated (Table 2).

The model’s C-statistic was 0.816 (95% CI, 0.774−0.859), and the bias-corrected C-statistic from bootstrapping was 0.807 (95% CI, 0.763−0.851). Calibration was demonstrated by the Hosmer−Lemeshow test (χ˄2_8 = 8.4279, p = 0.2964), indicating a good fit to the observed data.

A simple scoring system, CCDS65, was created from the β-coefficients (Table 3), assigning one point to each of the following variables: age between 18−64 years, male sex, history of CAD, history of CHF, and the presence of discordant LBBB. The C-statistics of this scoring system was 0.810 (95% CI, 0.767−0.853). The diagnostic performance of the scoring system in the development cohort—covering sensitivity, specificity, negative and positive likelihood ratios, and true positive and negative rates—is presented in Supplementary Table 3. Patients were subsequently stratified into four risk categories for developing reduced LV systolic function in patients with LBBB: very low probability (score 0), low probability (score 1−2), medium probability (score 3−4), and high probability (score 5−6).

**Table 3.** The CCDS65 scoring system.

|  |  |
| --- | --- |
| CAD history | 1 |
| CHF history | 2 |
| Discordant LBBB | 1 |
| Sex, males | 1 |
| Age, 18–64 years | 1 |

CHF, congestive heart failure; LBBB, left bundle branch block; CAD, coronary artery disease

### External validation of the model

In the validation cohort, the C-statistic (area under the curve) of the prediction model was recorded at 0.813 (95% CI, 0.772−0.855), and for the scoring system, it was 0.7806 (95% CI, 0.7367−0.8246). The diagnostic performance of the scoring system in the development cohort—including sensitivity, specificity, negative likelihood ratio, positive likelihood ratio, true positive, true negative, false negative, and false positive rates—are presented in Supplementary Table 4.

The mean predicted probabilities stratified by the CCDS65 scoring system showed varying risk levels: very low probability (score 0) at 3.27%, low probability (scores 1−2) at 11.78%, medium probability (scores 3−4) at 40.33%, and high probability (scores 5−6) at 73.34% (Figure 3). The predicted probabilities with 95% confidence intervals and the observed outcomes in both the development and validation cohorts are detailed in Figure 3. The correlation between predicted probabilities and observed outcomes for reduced LV systolic function in both cohorts confirmed the model’s accuracy and utility in predicting the presence of reduced LV systolic function in patients with LBBB.

**Figure 3.**
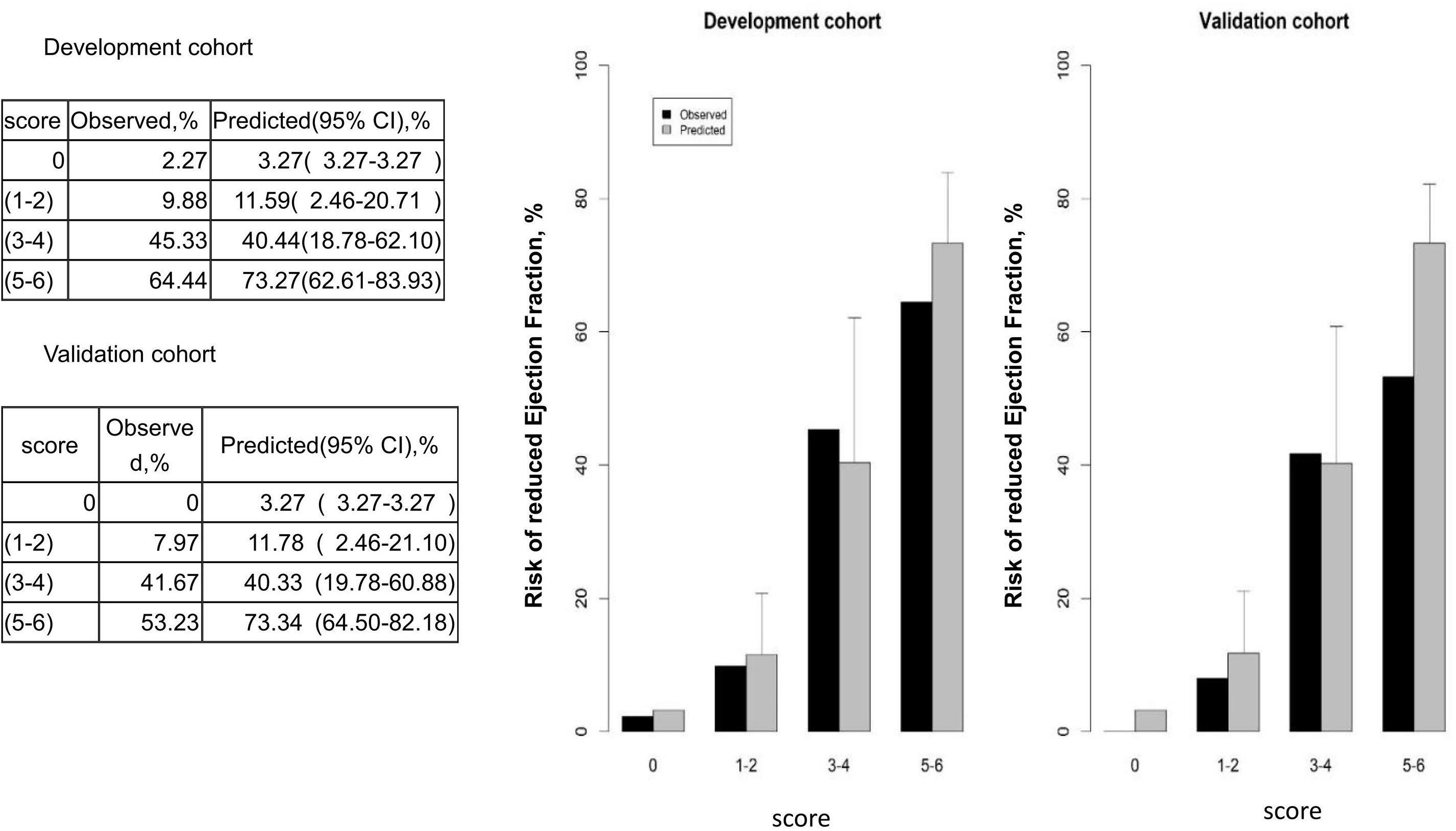
Predicted probability and observed risk of reduced ejection fraction (EF) in the development and validation cohorts.

## Discussion

Our study presents a simple yet robust predictive model for identifying reduced LV systolic function in patients with LBBB. With a promising AUC, this model utilized a 12-lead ECG and patient history to effectively stratify individuals at high risk for reduced LV systolic function. It offers a valuable tool for managing CAD and HF risk factors and tailoring follow-up frequency in clinical practice, thereby improving awareness and management of LBBB-induced cardiomyopathy. Although LBBB is relatively rare in the general population and the concurrent prevalence of complete LBBB with heart failure with reduced ejection fraction (HFrEF) is low (7), the incidence of patients developing LBBB-induced cardiomyopathy (LBBB-CM) within 4 years following an initial diagnosis of preserved LV function and LBBB is significant (17−38%) (8) (9). This condition is associated with poor outcomes, including higher mortality rates, reduced LVEF, and increased need for an implantable cardioverter-defibrillator compared to those without LBBB (10). Identifying patients who are at risk has not yet been addressed.

Previous research suggests that at the time of initial diagnosis of LBBB, patients with LVEF <60% and a left ventricular end-systolic diameter >29 mm are at a heightened risk of developing reduced LV systolic function (9) Additionally, another study identified diastolic filling time—constituting <38% of the cardiac cycle—as a significant predictor of reduced LV systolic function, with a relative risk of 7.0 [95% CI: 3.0−16.0]. (11) However, these findings were derived from cohorts with relatively small sample sizes.

This cross-sectional study assessed the near-term risk of reduced LV systolic function upon LBBB diagnosis, featuring a larger sample size (Total 821 patients, 229 patients with reduced LV systolic function), which enabled the creation of developmental and validation cohorts. The model considers established risk factors such as male sex, CAD, and CHF and includes younger patients and those with discordant LBBB electrocardiogram morphology. Our findings reinforce the role of discordant LBBB as an independent predictor of a higher risk for cardiovascular death or non-fatal heart failure hospitalization in patients with LBBB and an EF of ≥40%, underscoring the critical nature of these indicators in the early detection and management of heart disease (12).

The ability to screen patients at risk for LBBB with reduced LV systolic function has been complemented by emerging evidence supporting new pacing technologies (13–15) and advanced guideline-directed medical therapies (GDMT) (16). Recent advancements include early left bundle branch pacing combined with GDMT, which has been shown to significantly enhance cardiac function and reduce clinical events compared to GDMT alone in patients with heart failure with mildly to moderately reduced ejection fraction (HFmrEF, i.e., LVEF 36−50%) and LBBB (17). Furthermore, 12-month follow-ups have observed LV remodeling in patients with heart failure with preserved ejection fraction (HFpEF), although the effects were less pronounced than in HfmrEF (18). A pivotal pilot study conducted in 2024 applied cardiac resynchronization therapy to this patient group, demonstrating substantial improvements in LVEF and ventricular remodeling after 6 months (19). These findings demonstrate the potential benefits of early device intervention in patients with HFmrEF and LBBB, suggesting that such approaches may be crucial for LV remodeling. Identifying patients with LBBB and reduced LV systolic function is therefore imperative to leverage these therapeutic advancements effectively.

This study had several limitations. First, there is a potential risk of selection bias due to the hospital-based nature of our cohort, which might have influenced the awareness and reporting of disease status compared to a community-based sample. Second, although the size of our patient cohort was larger than those in previous studies of LBBB-CM predictors, the numbers were still relatively small. Third, our exclusion of patients with pacing-induced LBBB might limit the applicability of our model. Fourth, while our model predicts the near-term possibility of reduced LV systolic function upon LBBB diagnosis, it does not capture longitudinal data concerning the progression from LBBB diagnosis to the development of LBBB-CM. Finally, while internal validation within our cohort was performed, additional external validation is necessary to confirm the model’s accuracy across different populations.

## Conclusion

In conclusion, we successfully developed the CCDS65 scoring system, which effectively predicts echocardiographic reduced LV systolic function in patients with complete LBBB simply through history taking and ECG morphology. Given the advancements in GDMT and left bundle branch area pacing, early detection of reduced LV systolic function in patients with complete LBBB is increasingly important. Future research is essential to evaluate the scoring system’s performance in diverse cohorts, aiming to enhance its clinical accuracy and broaden its applicability.

## Non-standard Abbreviations and Acronyms

CAD: coronary artery disease
CHF: congestive heart failure
ECG: electrocardiogram
GDMT: guideline-directed medical therapies
HF: heart failure
HFrEF: heart failure with reduced ejection fraction
HfmEF: heart failure with mildly to moderately reduced ejection fraction
LBBB: left bundle branch block
LBBB-CM: left bundle branch block-induced cardiomyopathy
LV: left ventricular
LVEF: left ventricular ejection fraction
NTUH: National Taiwan University Hospital

## Data Availability

The dataset used and analyzed during the current study are available from the corresponding author on reasonable request.

## Acknowledgments

Chen CK, Liu YB, and Huang HC contributed to study conception and design. Chen CK and Huang HC contributed to the acquisition of the data. Chen CK, Liu YB, Chen CC, Chien KL, and Huang HC contributed to the analysis and/or interpretation of data. All authors contributed to the drafting or critical revision of the article for important intellectual content. All authors gave final approval of the version of the article to be published. The authors also thank Wiley Editing Services for English language editing.

## Supplemental Material

Tables S1−S4

## References

1. Sze E, Samad Z, Dunning A, Campbell KB, Loring Z, Atwater BD, Chiswell K, Kisslo JA, Velazquez EJ, Daubert JP Impaired Recovery of Left Ventricular Function in Patients With Cardiomyopathy and Left Bundle Branch Block. J Am Coll Cardiol 2018;71:306–317.

2. Sze E, Daubert JP. Left bundle branch block-induced left ventricular remodeling and its potential for reverse remodeling. Journal of Interventional Cardiac Electrophysiology: An International Journal of Arrhythmias and Pacing. 2018;52:343–352.

3. Huizar JF, Kaszala K, Tan A, Koneru J, Mankad P, Kron J, Ellenbogen KA. Abnormal Conduction-Induced Cardiomyopathy: JACC Review Topic of the Week. Journal of the American College of Cardiology. 2023;81:1192–1200.

4. Collins GS, Reitsma JB, Altman DG, Moons KG. Transparent reporting of a multivariable prediction model for individual prognosis or diagnosis (TRIPOD): the TRIPOD statement. BMJ (Clinical Research Ed.). 2015;350:g7594.

5. Surawicz B, Childers R, Deal BJ, Gettes LS, Bailey JJ, Gorgels A, Hancock EW, Josephson M, Kligfield P, Kors JA, et al. AHA/ACCF/HRS recommendations for the standardization and interpretation of the electrocardiogram: part III: intraventricular conduction disturbances: a scientific statement from the American Heart Association Electrocardiography and Arrhythmias Committee, Council on Clinical Cardiology; the American College of Cardiology Foundation; and the Heart Rhythm Society. Endorsed by the International Society for Computerized Electrocardiology. Journal of the American College of Cardiology. 2009;53:976–981.

6. Mitchell C, Rahko PS, Blauwet LA, Canaday B, Finstuen JA, Foster MC, Horton K, Ogunyankin KO, Palma RA, Velazquez EJ. Guidelines for performing a comprehensive transthoracic echocardiographic examination in adults: Recommendations from the American Society of Echocardiography. Journal of the American Society of Echocardiography: Official publication of the American Society of Echocardiography. 2019;32:1–64.

7. Huang HC, Chien KL, Lin HJ, Liu YB. The prevalence and association of patients with impaired left ventricular ejection fraction and complete left bundle-branch block in Taiwan. Journal of the Formosan Medical Association = Taiwan yi zhi. 2019;118:686–691.

8. Lee SJ, McCulloch C, Mangat I, Foster E, De Marco T, Saxon LA. Isolated bundle branch block and left ventricular dysfunction. Journal of Cardiac Failure. 2003;9:87–92.

9. Sharma S, Barot HV, Schwartzman AD, Ganatra S, Shah SP, Venesy DM, Patten RD. Risk and predictors of dyssynchrony cardiomyopathy in left bundle branch block with preserved left ventricular ejection fraction. Clinical Cardiology. 2020;43:1494–1500.

10. Witt CM, Wu G, Yang D, Hodge DO, Roger VL, Cha YM. Outcomes with left bundle branch block and mildly to moderately reduced left ventricular function. JACC. Heart Failure. 2016;4:897–903.

11. Atwater BD, Emerek K, Samad Z, Sze E, Black-Maier E, Loring Z, Ugander M, Liao L, Kisslo J, Søgaard P, et al. Predicting the development of reduced left ventricular ejection fraction in patients with left bundle branch block. The American Journal of Cardiology. 2020;137:39–44.

12. Huang HC, Chien KL, Liu YB. Outcomes with T-wave discordance of left bundle branch block and preserved or mildly reduced ejection fraction. ESC Heart Failure. 2024;11:2148–2158.

13. Cheng L, Zhang J, Wang Z, Zhou M, Liang Z, Zhao L, Chen J, Wu Y. Efficacy and safety of left bundle branch area pacing versus biventricular pacing in heart failure patients with left bundle branch block: study protocol for a randomised controlled trial. BMJ Open. 2020;10:e036972.

14. Chen X, Ye Y, Wang Z, Jin Q, Qiu Z, Wang J, Qin S, Bai J, Wang W, Liang Y, et al. Cardiac resynchronization therapy via left bundle branch pacing vs. optimized biventricular pacing with adaptive algorithm in heart failure with left bundle branch block: a prospective, multi-centre, observational study. Europace: European pacing, arrhythmias, and cardiac electrophysiology: journal of the working groups on cardiac pacing, arrhythmias, and cardiac cellular electrophysiology of the European Society of Cardiology. 2022;24:807–816.

15. Ponnusamy SS, Vijayaraman P. Left bundle branch block-induced cardiomyopathy: insights from left bundle branch pacing. JACC. Clinical Electrophysiology. 2021;7:1155–1165.

16. Wang NC, Singh M, Adelstein EC, Jain SK, Mendenhall GS, Shalaby AA, Voigt AH, Saba S. New-onset left bundle branch block-associated idiopathic nonischemic cardiomyopathy and left ventricular ejection fraction response to guideline-directed therapies: The NEOLITH study. Heart Rhythm. 2016;13:933–942.

17. Zeng J, He C, Zou F, Qin C, Xue S, Zhu H, Li X, Liu Z, Wei Y, Hou S, et al. Early left bundle branch pacing in heart failure with mildly reduced ejection fraction and left bundle branch block. Heart Rhythm. 2023;20:1436–1444.

18. Ye Y, Chen X, He L, Wu S, Su L, He J, Zhang Y, Sheng X, Yu C, Yang Y, et al. Left bundle branch pacing for heart failure and left bundle branch block patients with mildly reduced and preserved left ventricular ejection fraction. The Canadian Journal of Cardiology. 2023;39:1598–1607.

19. Cha YM, Lee HC, Mulpuru SK, Deshmukh AJ, Friedman PA, Asirvatham SJ, Bradley DJ, Madhavan M, Abou Ezzeddine OF, Wen S, et al. Cardiac resynchronization therapy for patients with mild to moderately reduced ejection fraction and left bundle branch block. Heart Rhythm. 2024;21:2250–2259.

